# Role of non-classical monocytes in HIV-associated vascular cognitive impairment

**DOI:** 10.1101/2023.03.24.23287660

**Authors:** Meera V Singh, Md Nasir Uddin, Mae Covacevich Vidalle, Karli R. Sutton, Zachary D. Boodoo, Angelique N. Peterson, Alicia Tyrell, Raeann Brenner, Madalina E Tivarus, Henry Z. Wang, Bogachan Sahin, Jianhui Zhong, Miriam Weber, Lu Wang, Xing Qiu, Sanjay B. Maggiwar, Giovanni Schifitto

## Abstract

Despite antiretroviral treatment (cART), people living with HIV (PLWH) are more susceptible to neurocognitive impairment (NCI), probably due to synergistic/additive contribution of traditional cerebrovascular risk factors. Specifically, altered blood brain barrier (BBB) and transmigration of inflammatory monocytes are risk factors for developing cerebral small vessel disease (CSVD). In order to investigate if inflammatory monocytes exacerbate CSVD and cognitive impairment, 110 PLWH on cART and 110 age-, sex- and Reynold’s cardiovascular risk score-matched uninfected individuals were enrolled. Neuropsychological testing, brain magnetic resonance imaging and whole blood analyses to measure platelet-monocyte interaction and monocyte, endothelial activation were performed. Results demonstrated that PLWH exhibited increased levels of platelet-monocyte complexes (PMCs) and higher expression of activation molecules on PMCs. PLWH with CSVD had the poorest cognitive performance and the highest circulating levels of non-classical monocytes which exhibited significant inverse correlation with each other. Furthermore, markers of monocyte and endothelium activation were significantly positively correlated indicating BBB impairment. Our results confirm that interaction with platelets activates and drives monocytes towards an inflammatory phenotype in PLWH. In particular, elevated levels of non-classical monocytes may represent a common pathway to neuroinflammation, CSVD and subsequent cognitive impairment, warranting further longitudinal studies to evaluate responsiveness of this potential biomarker.

## INTRODUCTION

HIV associated, age-related comorbidities have a taken a center-stage in the management of this disease, owing to the increased longevity of people living with HIV (PLWH) in the post combination anti-retroviral therapy (cART) era. Among these, cerebral small vessel disease (CSVD) is one of the most enigmatic vascular diseases, which is known to be more prevalent in PLWH than the general population, as reported by our group (1) and others (2–5). CSVD affects small penetrating arteries, arterioles, capillaries and small veins. It covers a range of pathological and clinical abnormalities such as small subcortical infarcts, lacunes, white matter hyperintensities (WMH), enlarged perivascular spaces (EPVS), cerebral microbleeds (CMB) and brain atrophy that can only be detected by neuroimaging techniques (6, 7) thus making its diagnosis, which is based only on clinical examination, technically challenging. CSVD is considered to be a major contributor to vascular cognitive impairment (VCI) in the older population (2, 8, 9). Currently, asymptomatic and mild neurocognitive impairment (ANI and MND) are the most prevalent forms of HIV associated neurological disorder (HAND). While not as severe as HIV-associated dementia (HAD), these are still clinically relevant because individuals with ANI are two to six times more likely to develop symptomatic HAND (10, 11). Studies in resource-limited settings such as sub-Saharan Africa, where the majority of PLWH reside, indicate that HAND might be the most prevalent form of neurocognitive impairment (NCI) (12) in this region. However, in the aging HIV population, the clinical phenotype of cognitive impairment includes the effects of chronic peripheral immune activation and inflammation (3, 5, 13) on brain vasculature.

In this regard, monocytes have a multifaceted role in HIV infection. There are three subsets of circulating monocytes defined by the difference in expression pattern for CD14 and CD16 as classical (CD14++CD16-), intermediate (CD14++CD16+) and non-classical (CD14+CD16++) monocytes. It has been suggested that monocytes sequentially mature from classical to intermediate to non-classical phenotype during circulation (14–16). CD16+ monocytes are considered to be responsible for seeding and maintenance of CNS viral reservoir as well as for increased neuroinflammation during HIV infection (17–20). Previous reports have shown that the CD16+ phenotype is also induced by the interaction of monocytes with activated platelets (21, 22). Activated platelets form transient complexes with monocytes in circulation (platelet-monocyte complexes, PMCs), and these complexes are elevated in diseases involving inflammation such as cardiovascular disease (23, 24). Previous studies from our group (25) and others (26) have demonstrated increased platelet activation during HIV infection. PLWH, especially those with VCI, have been shown to exhibit an increase in plasma and cerebrospinal fluid levels of sCD40L (27), for which platelets are the major source (28). Increased interaction between platelets and monocytes induces a proinflammatory, promigratory phenotype in monocytes and has significant implications for HIV associated neuroinflammation (22).

In order to further investigate the pathogenic role of platelet-monocyte interaction and subsequent monocyte activation in HIV-associated CSVD as well as VCI, 110 PLWH on stable cART for at least three months before enrollment and an equal number of age, sex-matched healthy individuals (29) were enrolled in this study. These individuals underwent comprehensive neuropsychological testing, brain MRI, whole blood analyses to characterize platelet-monocyte interaction and markers of monocyte activation on PMCs. Soluble products of monocyte (sCD14, CD163) and endothelial (ICAM, VCAM, Osteoprotegerin and lipoprotein associate phospholipase A2-LpPLA2) activation were also measured. These markers have been previously linked with CSVD in general population (30, 31) and also with development and progression of HIV-associated atherosclerosis (32–35). Based on the MRI results and HIV status, the participants were divided into 4 groups: PLWH with CSVD (HIV+CSVD+), PLWH without CSVD (HIV+CSVD), uninfected individuals with CSVD (HIV-CSVD+) and without CSVD (HIV-CSVD-).

Our results indicate that the combination of HIV and CSVD is most detrimental to neurocognition. Chronic inflammation during HIV infection exacerbates platelet-monocyte interaction and subsequent monocyte activation, especially of the non-classical monocyte subset. Levels of non-classical monocytes showed very strong inverse correlation with cognitive performance in individuals with both HIV and CSVD. Longitudinal studies in PLWH are warranted to evaluate the role of non-classical monocytes as a potential biomarker to monitor progression of CSVD and associated cognitive impairment.

## RESULTS

### HIV infected individuals with CSVD exhibit significant cognitive impairment

Results indicate that PLWH with CSVD (n=77) showed significantly lower total cognitive score compared to HIV uninfected individuals without CSVD (n=38, p=0.0002) and with CSVD (n=66, p=0.0001, **figure 1A**). Individuals with HIV and CSVD both also showed significantly reduced scores for all but one of the individual cognitive domains, including processing speed (**figure 1B**, p=0.0076 and 0.0052), executive function (**figure 1C**, p=0.0291 and 0.000.1), fine motor skills (**figure 1D**, p=0.003 and 0.0011) and verbal and visual learning (**figure 1E**, p=0.0012 and 0.0133) compared to both HIV uninfected groups. Within the HIV+ groups, those with CSVD had significantly lower scores in only one cognitive domain, fine motor skills, compared to the HIV+CSVD-group (**figures 1D**, p=0.0089). Individuals that were HIV+CSVD-had significantly lower verbal and visual learning scores compared to both HIV uninfected groups (**figure 1E**, p=0.0176 and 0.0047). Both HIV+ groups had significantly lower language scores compared to HIV-CSVD+ group (**figure 1F**, p=0.0419, 0.0042), while only the HIV+CSVD+ group had a significantly lower verbal and visual memory score compared with the HIV-CSVD+ group (**figure 1G**, p=0.0352). Lastly, the four groups did not differ in the attention and working memory score (**figure 1H**).

**Figure 1:**
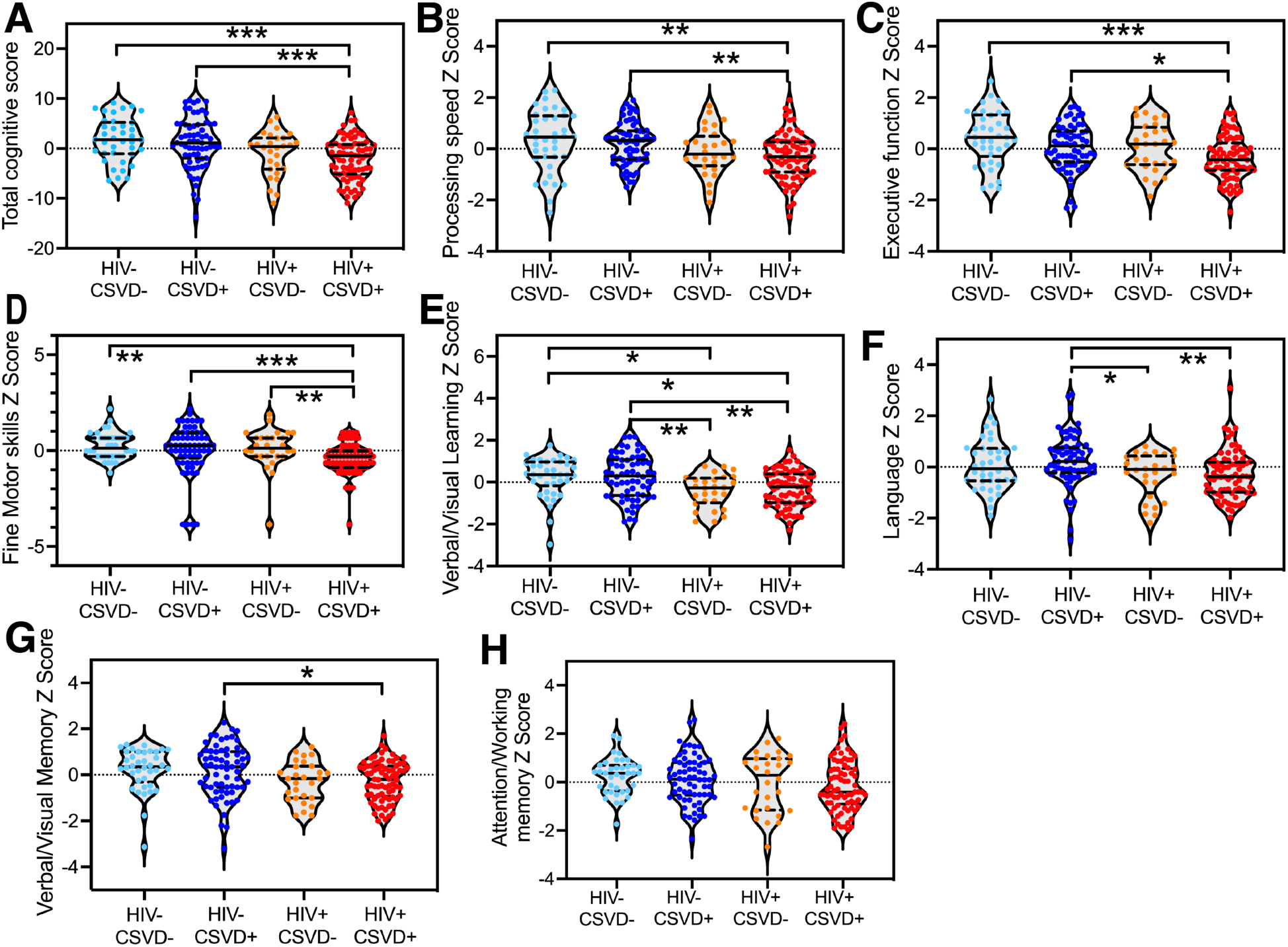
Neuropsychological battery domain scores in study participants. **A.** Total cognitive score. Individual domain scores for **B.** Processing speed, **C.** Executive function, **D.** Fine Motor Skill, **E.** Verbal and Visual Learning, **F.** Language, **G.** Verbal and Visual Memory and, **H.** Attention and working memory. Light blue data points represent HIV-CSVD-group (n=38), dark blue represent HIV-CSVD+ group (n=66), orange represent HIV+CSVD-group (n=28) and red represent HIV+CSVD+ group (n=77). Four group comparisons by Kruskal-Wallis test followed by Dunn’s multiple comparisons test. correlation analysis by Spearman’s rank correlation test. * p<0.05, ** p<0.01, *** p<0.001.

### Increased levels of platelet activation, platelet-monocyte complexes (PMCs) and non-classical monocytes were found in PLWH

Percentages of CD62p+ platelets were measured as a marker of platelet activation in whole blood by flow cytometry. Results showed that platelets were more activated in PLWH (**Figure 2A**, p=0.0024, HIV-n=84, HIV+ n=97). Additional categorization based on CSVD status showed that the HIV+CSVD+ group had the highest levels of platelet activation among the 4 groups and this increase was statistically significant as compared to HIV-CSVD-individuals (**Figure 2B**, p=0.0123, HIV-CSVD-n=31, HIV-CSVD+ n=53, HIV+CSVD-n=26, HIV+CSVD+ n=71). Next, levels of different monocyte subsets and platelet-monocyte complexes within each monocyte subset were measured by flow cytometry using whole blood. Monocytes were divided into three subsets based on the expression of CD14 and CD16 markers as, classical monocytes (CD14^+^CD16^-^), intermediate monocytes (CD14^+^CD16^+^) and non-classical monocytes (CD14^lo^CD16^+^). Results indicated that levels of circulating PMCs are significantly increased among all three monocyte subsets in PLWH compared to HIV-individuals (**Figure 2C**, p=0.0006 for non-classical PMCs, **supplementary Figure 1A**, p=0.0002 for classical PMCs, **supplementary Figure 1B**, p=0.0003 for intermediate PMCs, HIV-n=81, HIV+ n=94). This increase in PMCs among all three monocyte subsets was mostly due to HIV status rather than CSVD status (**supplementary Figures 1C-1E,** HIV-CSVD-n=31, HIV-CSVD+ n=50, HIV+CSVD-n=26, HIV+CSVD+ n=68). Higher levels of platelet activation correlated positively with increased levels of PMCs within all three monocyte subsets (**Figure 2D**, p=0.0002, r=0.2842, for non-classical PMCs; n=172, data not shown for other subsets). PLWH also showed a significant increase in the levels of non-classical monocytes (**Figure 2E**, p<0.0001, HIV-n=81, HIV+ n=94). Classical monocytes showed a trend towards decreased levels in PLWH (**supplementary Figure 1F,** p=0.056, HIV-n=81, HIV+ n=94), but there was no difference in the levels of intermediate monocytes (**supplementary Figure 1G,** HIV- n=81, HIV+ n=94). Interestingly, when categorized based on CSVD and HIV status, highest levels of non-classical monocytes were found in HIV+CSVD+ as compared with HIV-CSVD- individuals (p=0.0011) as well as HIV-CSVD+ individuals (p=0.0003, **Figure 2F**, HIV-CSVD- n=31, HIV-CSVD+ n=50, HIV+CSVD- n=26, HIV+CSVD+ n=68), indicating that HIV and CSVD both are associated with higher levels on non-classical monocytes.

**Figure 2:**
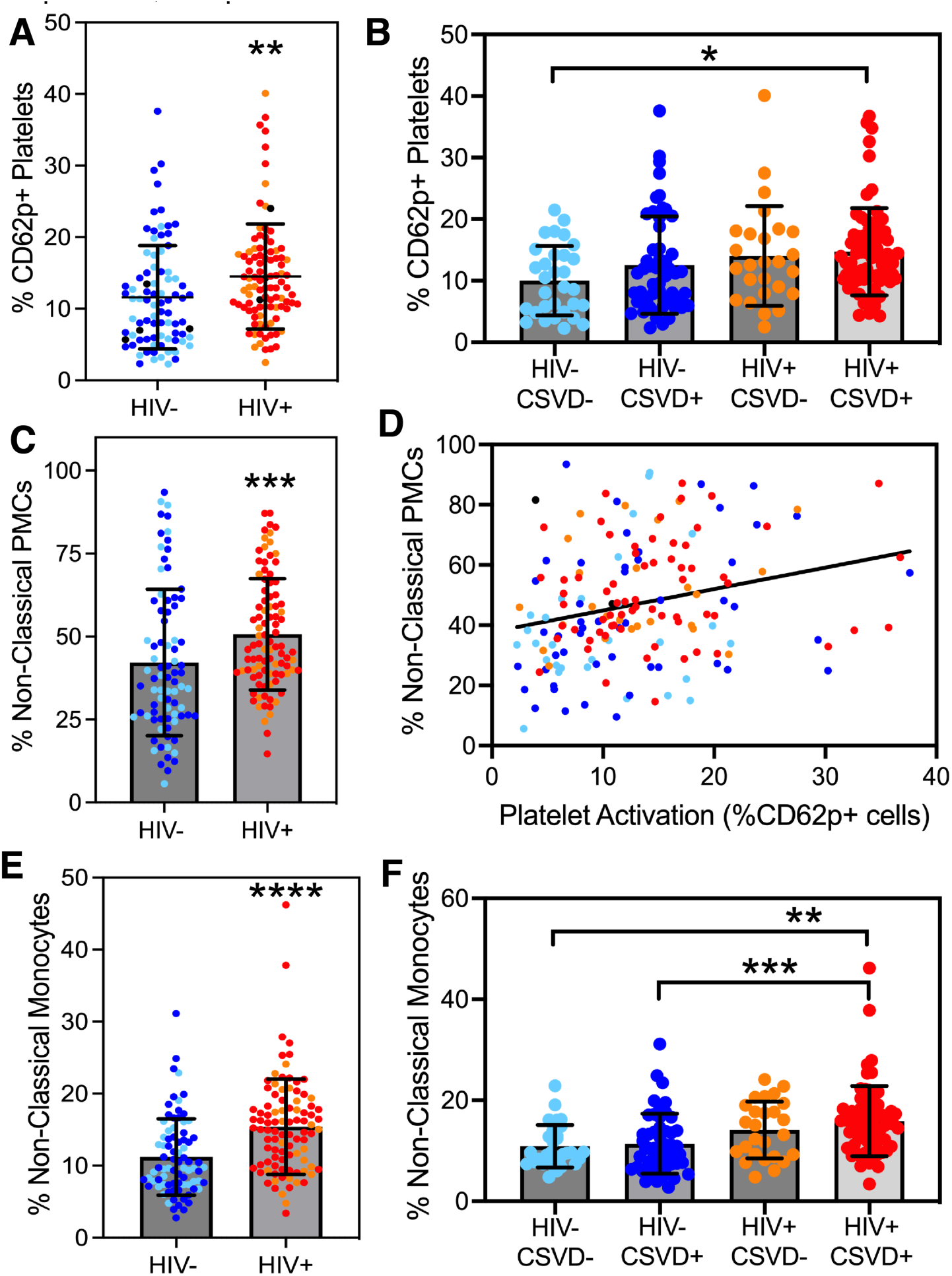
Flow cytometric analysis of platelets activation, platelet-monocyte interaction. Percentages of platelets expressing of CD62p, a marker of platelet activation, **A.** by HIV status (HIV- n=84, HIV+ n=97), **B.** by HIV and CSVD status (HIV-CSVD- n=31, HIV-CSVD+ n=53, HIV+CSVD- n=26, HIV+CSVD+ n=71). **C.** Percentages of platelet-monocyte complexes (PMCs) among all non-classical monocytes (HIV- n=81, HIV+ n=94). **D.** Correlation analysis between percentages of non-classical PMCs and CD62p+ platelets (n=172, r=0.2842, p=0.0002). Percentages of non-classical monocytes (irrespective of whether the cells are complexed to platelets or not) categorized based on, **E.** HIV status (HIV- n=81, HIV+ n=94) or **F.** HIV and CSVD status (HIV-CSVD- n=31, HIV-CSVD+ n=50, HIV+CSVD- n=26, HIV+CSVD+ n=68). Light blue data points represent HIV-CSVD- group, dark blue represent HIV-CSVD+ group, orange represent HIV+CSVD- group and red represent HIV+CSVD+ group. Two group comparisons by Mann-Whitney U test, four group comparisons by Kruskal-Wallis test followed by Dunn’s multiple comparisons test. correlation analysis by Spearman’s rank correlation test. * p<0.05, ** p<0.01, *** p<0.001, **** p<0.0001.

### Elevated expression of monocyte activation markers based on HIV status and platelet interaction

Expression of monocyte activation markers CCR2, CD40, PSGL-1, TNFR2 and TF was measured on monocytes as well as PMCs by flow cytometry. Percentages of non-classical (**Figure 3A**, p=0.0172, HIV- n=80, HIV+ n=93), classical and intermediate monocytes (**supplementary Figure 2A and 2B**, p=0.0149, p=0.0075 respectively, HIV- n=80, HIV+ n=93) expressing tissue factor (TF) were found to be increased in PLWH as compared to uninfected individuals. Classical monocytes also showed increased expression of CD40 in PLWH (**supplementary Figure 2C**, p=0.0077, HIV- n=81, HIV+ n=94). There was no difference between the two groups with respect to other markers. Notably, non-classical PMCs showed increased percentage of cells expressing TF (n=173), CD40 (n=175), TNFR2 (n=175) and CCR2 (n=174) as compared to non-complexed monocytes (non-PMCs) irrespective of their HIV status (**Figure 3B-3E**, p<0.0001 for all markers). PSGL-1 did not show any difference between non-classical PMCs and non-PMCs (**Figure 3F**, n=175). Classical PMCs showed significantly increased percentage of cells expressing TF (n=173), CD40 (n=175) and TNFR2 (n=175) as compared to non-PMCs (**supplementary Figures 2D-2F,** p<0.0001 for all markers). Intermediate PMCs showed increased expression of TF only (**supplementary Figure 2G,** p<0.0001, n=175).

**Figure 3:**
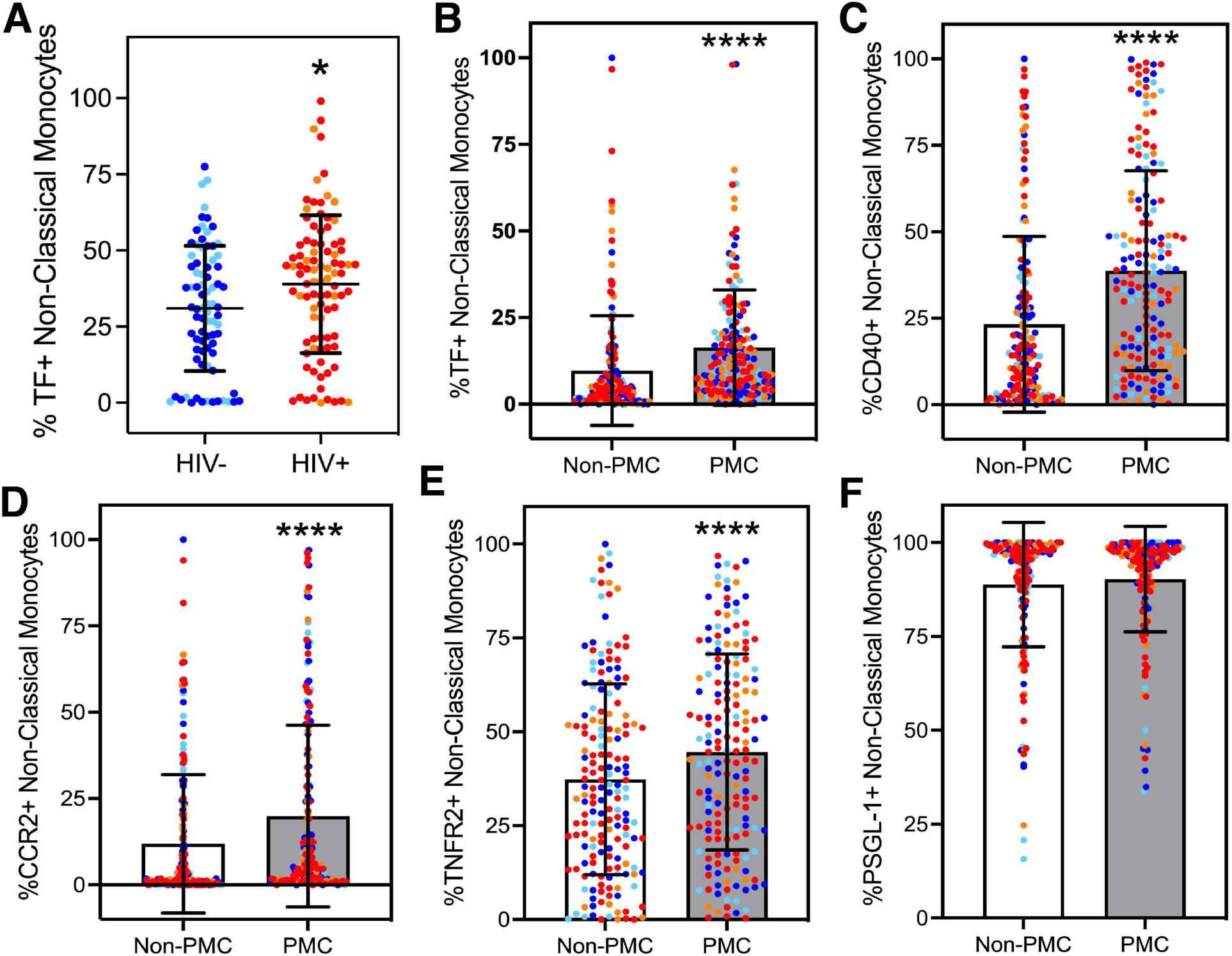
Flow cytometric analysis of monocyte activation for non-classical subset of monocytes. **A.** Levels of non-classical monocytes expressing tissue factor (irrespective of whether the cells are complexed to platelets or not, HIV- n=80, HIV+ n=93). Two group comparisons by Mann-Whitney U test, * p<0.05. Comparison between percentages of platelet- complexed (PMCs) and non-complexed (non-PMCs) non-classical monocytes for expression levels of **B.** Tissue factor (TF, n=173), **C**. CD40 (n=175), **D.** Chemokine receptor 2 (CCR2, n=174), **E.** Tissue necrosis factor receptor 2 (TNFR2, n=175), **F.** p-selectin glycoprotein ligand-1 (PSGL-1, n=175). Light blue data points represent HIV-CSVD- group, dark blue represent HIV- CSVD+ group, orange represent HIV+CSVD- group and red represent HIV+CSVD+ group. Two group comparisons between PMCs and non-PMCs by Wilcoxon signed-rank test, **** p<0.0001.

### Soluble ICAM and CD14 levels are increased in HIV infected individuals with CSVD

Plasma levels of endothelial activation markers ICAM, VCAM, Osteoprotegerin and Lp- PLA2 as well as monocyte/macrophage activation marker CD163 and sCD14 were measured by ELISA assays. ICAM levels were found to be increased in HIV infected individuals (p=0.0093, **Figure 4A**, HIV- n=104, HIV+ n=105). When categorized based on HIV and CSVD status, individuals with HIV and CSVD showed significantly higher ICAM levels as compared to HIV uninfected individuals with CSVD (p=0.0295, **Figure 4B**, HIV-CSVD- n=39, HIV-CSVD+ n=66, HIV+CSVD- n=27, HIV+CSVD+ n=77). There were no statistically significant differences observed in the levels of VCAM (HIV-CSVD- n=37, HIV-CSVD+ n=65, HIV+CSVD- n=26, HIV+CSVD+ n=78), Lp-PLA2 (HIV-CSVD- n=39, HIV-CSVD+ n=65, HIV+CSVD- n=28, HIV+CSVD+ n=79), and Osteoprotegerin (HIV-CSVD- n=37, HIV-CSVD+ n=65, HIV+CSVD-n=27, HIV+CSVD+ n=77) (**supplementary Figures 3A-3C**). Among the monocyte/macrophage soluble markers, sCD14 was found to be significantly increased in PLWH (p=0.0001, **Figure 4C**, HIV- n=105, HIV+ n=106) with PLWH with CSVD exhibiting the highest levels as compared to both the HIV uninfected groups, with and without CSVD (p=0.0283 and p-0.0208 respectively, **Figure 4D**, HIV-CSVD- n=40, HIV-CSVD+ n=65, HIV+CSVD- n=28, HIV+CSVD+ n=78). While statistically similar levels of CD163 were noted in the plasma of HIV+ and HIV- groups (**supplementary Figure 3D,** HIV- n=103, HIV+ n=105), individuals that were HIV+CSVD- showed significantly increased levels of CD163 as compared with HIV-CSVD- group (p=0.0444, **supplementary Figure 3E,** HIV-CSVD- n=40, HIV-CSVD+ n=63, HIV+CSVD- n=27, HIV+CSVD+ n=78). Interestingly, ICAM and sCD14 levels were highly positively correlated with each other (r=0.4285, p<0.0001, **Figure 4E**, n=206).

**Figure 4:**
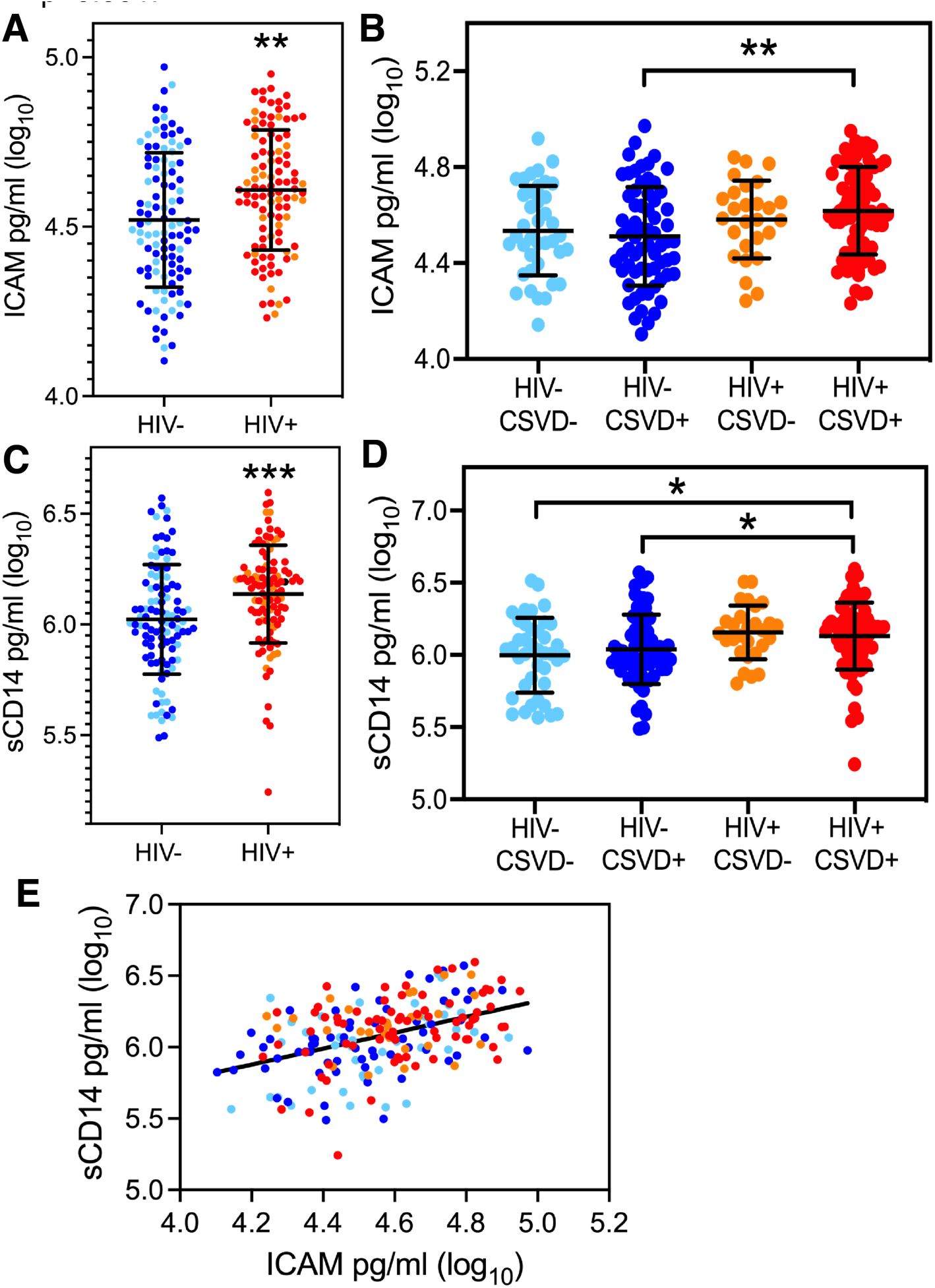
Soluble markers of endothelial and monocyte activation. Plasma levels of intracellular adhesion molecule (ICAM) in study participants when categorized based on **A.** HIV status (HIV- n=104, HIV+ n=105) and **B.** HIV and CSVD status (HIV-CSVD- n=39, HIV-CSVD+ n=66, HIV+CSVD- n=27, HIV+CSVD+ n=77). Plasma levels of soluble CD14 (sCD14) in study participants when categorized based on **C.** HIV status (HIV- n=105, HIV+ n=106) and **D.** HIV and CSVD status (HIV-CSVD- n=40, HIV-CSVD+ n=65, HIV+CSVD- n=28, HIV+CSVD+ n=78). **E.** Correlation analysis between ICAM and sCD14 (n=206, r= 0.4285, p<0.0001). Light blue data points represent HIV-CSVD- group, dark blue represent HIV-CSVD+ group, orange represent HIV+CSVD- group and red represent HIV+CSVD+ group. Two group comparisons by Mann- Whitney U test, four group comparisons by Kruskal-Wallis test followed by Dunn’s multiple comparisons test. Correlation analysis by Spearman’s rank correlation test. * p<0.05, ** p<0.01, *** p<0.001.

### Circulating levels of non-classical monocytes might be predictive of cognitive performance in HIV-associated CSVD

All three subsets of monocytes and PMCs as well as soluble markers of endothelial and monocyte activation were assessed against cognitive scores (total and individual domain scores). Results showed that when values from HIV+ and HIV- groups were pooled together, a negative correlation was observed between percentages of non-classical monocytes and total cognitive score (r=-2334, p=0.0021, **Figure 5A**, n=172). When categorized based on HIV status, there was no significant correlation among these two parameters in the HIV uninfected individuals (r=0.03068, **supplementary Figure 3F,** n=80), however the correlation was found to be stronger in PLWH (r=-0.2858, p=0.0058, **Figure 5B**, n=92). Interestingly, even among PLWH, it was the individuals with CSVD that seemed to drive the negative correlation between non-classical monocytes and total cognitive score (r=-0.3579, p=0.0032, **Figure 5C**, n=66) and the HIV+CSVD- group showed no correlation (r=-0.0906, **supplementary Figure 3G,** n=26). Similar negative correlations were found between percentages of non-classical monocytes and individual cognitive domain scores (summarized in **Table 2**, only statistically significant associations are shown). Soluble ICAM (r=-0.1533, p=0.027 **Figure 5D**, n=207) and Osteoprotegerin (r=-0.1842, p=0.009. **Figure 5E**, n=206) levels exhibited a mild negative correlation with total cognitive score when all data points from HIV+ and HIV- groups were pooled together. However, there were no correlations between ICAM, Osteoprotegerin and total cognitive score when values were categorized based on HIV status, or CSVD status alone or a combination of HIV and CSVD status.

**Figure 5:**
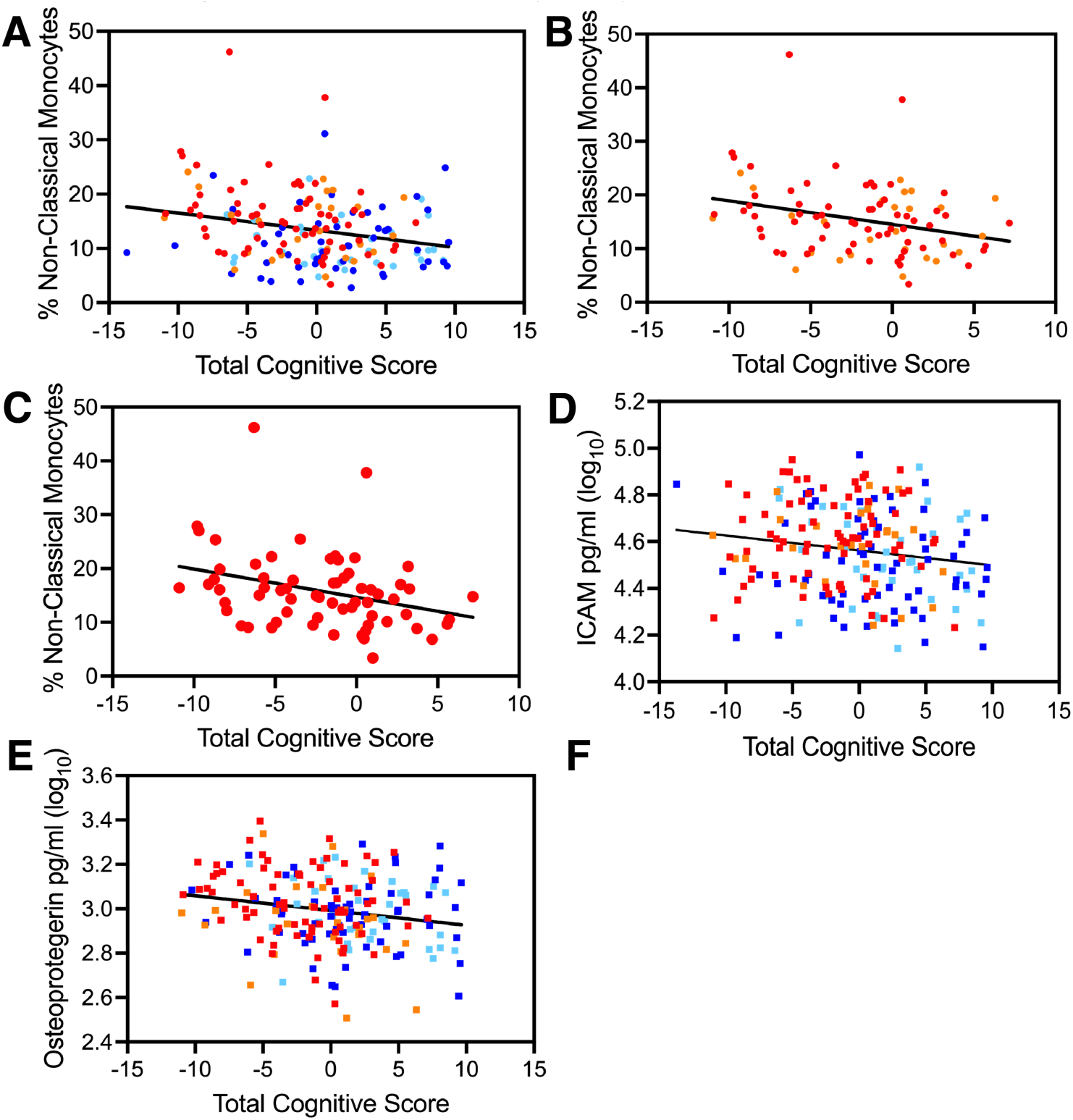
Correlation analysis between peripheral markers and cognitive performance. Correlation analysis between percentages of non-classical monocytes and total cognitive score **A.** when data from all study participants is pooled together (n=172, r=-0.2334, p=0.0021), **B.** HIV+ study group (n=92, r=-0.2858, p=0.0058), and **C.** HIV+CSVD+ study group (n=66, r=-0.3579, p=0.0032). Correlation analysis between **D.** ICAM (n=207, r=-0.1487, p=0.0286) or **E.** Osteoprotegerin (n=206, r=-0.1823, p=0.0087) and total cognitive score when data from all study participants is pooled together. Light blue data points represent HIV-CSVD- group, dark blue represent HIV-CSVD+ group, orange represent HIV+CSVD- group and red represent HIV+CSVD+ group. Correlation analysis by Spearman’s rank correlation test.

Further, we correlated the blood markers and total cognitive score with absolute WMH volumes (log transformed), however, no significant correlations between these measures were found. It should be noted that most scores for CSVD were between 1 and 2 indicating that the volume of WMH was relatively small.

## DISCUSSION

This study was designed to investigate the hypothesis that interaction with activated platelets inculcates a proinflammatory, promigratory phenotype in monocytes, which upon extravasation across BBB could exacerbate CSVD and vascular cognitive impairment in PLWH. Results from neuropsychological assessment indicate that while HIV and CSVD were both individually associated with cognitive impairment, a combination of the two was most detrimental to cognition. While early initiation of cART is known to improve cognition to some extent (36), we have recently shown that over time, PLWH relapse into worsening cognition as compared to uninfected individuals (37). Our current findings suggest that the presence of CSVD in PLWH seems to confer a “double hit” on cognitive function and might not be the exclusive mechanism of NCI in HIV infection. Rather, inflammatory, non-classical monocytes may play a key role in pathomechanisms of HIV-associated cognitive impairment and CSVD.

In this context, investigations on platelet-monocyte interaction in circulation showed that higher platelet activation in PLWH, especially those with CSVD, leads to excessive monocyte activation as compared to HIV- group. In addition, higher expression of monocyte activation markers (CCR2, CD40, TF and TNFR2) was present on PMCs as compared to non-PMCs. Further, increased levels of TF expression were observed on monocytes from PLWH, irrespective of whether they were complexed to platelets or not, suggesting that they might be more coagulopathic in nature. Several reports, including our group, have shown that PMCs (or aggregates) are elevated during cardiovascular disease, HIV and SARS-CoV-2 infection (21, 22, 24, 38–44). However, to the best of our knowledge, this is the first report that has conducted an in depth, ex vivo characterization of the expression of monocyte activation markers (PSGL-1, CCR2, CD40, TNFR2 and TF) on PMCs and shown that monocytes in complex with platelets express higher levels of these activation markers in whole blood. Monocytes hyperactivated in such a way might have a higher propensity to differentiate into inflammatory CD16+ non-classical subset, as was seen by the increase in their levels in circulation in the HIV+CSVD+ study group. In addition, a statistically significant inverse correlation was observed between percentages of non-classical monocytes and cognitive performance when data from all study participants was pooled together. Interestingly, this correlation became progressively stronger within just the HIV+ cohort followed by the most robust correlation coefficient for the HIV+CSVD+ group. These findings have significant implications with respect to neuroinflammation, because interaction with platelets is considered to confer a promigratory phenotype to monocytes (22, 45–47), and drive their differentiation to CD16+ monocytes, which upon entry into the CNS can cause excessive neuroinflammation. Indeed, Zuchtriegel et al found that, platelets spatiotemporally guide monocytes and are essential for effective trafficking of leukocytes to sites of inflammation (48).

Non-classical monocytes alternatively called patrolling monocytes are responsible for resolution of inflammation during homeostasis (49–51). However, multiple studies have also shown that these cells are equally capable of partaking in the pathogenesis of inflammatory diseases including vascular disease (reviewed in (52)) and are considered a heterogenous population. For example, recent studies have identified several other markers that can be used to further classify non-classical monocytes, including CD11c, CD36, CCR2, and CD163 (53–55). This heterogeneity of non-classical monocytes is thought to reflect the diversity of functions that they perform in the immune system. Increased CD16+ monocytes have been shown to be associated with severity of coronary artery disease (56) as well as be an independent predictor of cardiovascular events (57). Several reports have also shown either higher levels of circulating monocytes or soluble monocyte products to be elevated during cognitive decline associated with sepsis, neuropathic injury, Alzheimer’s disease and stress (58–63). With respect to HIV infection, which is considered a pro-atherosclerotic condition, non-classical monocytes were shown to predict the progression of carotid artery bifurcation intima media thickness (64). A study by Shikuma et al. showed that higher antiretroviral monocyte efficacy score was linked with better cognitive performance (65). HIV DNA levels in monocytes and soluble CD14 were found to be associated with VCI in PLWH (66, 67). Recently, Veenhuis et al showed that higher intermediate monocytes were associated with lower global neuropsychological function score in two small cohorts of 25 and 18 women respectively (68). However, the role of monocytes in CSVD, which is another form of vascular disease prevalent in PLWH, and also a major factor behind neurocognitive impairment, remains elusive. Only one report so far found that CD16+ monocytes were positively correlated with CSVD in a cohort of 51 HIV infected individuals (69) and we believe that our study is significant step towards bridging this gap in knowledge.

Another interesting insight into the interplay between periphery and neuronal inflammation was provided by assessment of soluble markers of monocyte and endothelial activation, ICAM and sCD14, which were found to be increased during HIV associated CSVD. Both of these markers positively correlated with each other very strongly, indicating that monocyte activation coincided with endothelial activation, a key component of the BBB. Lastly, ICAM, and osteoprotegerin correlated negatively with cognitive performance to some extent when all data points from each cohort were pooled together. However, contrary to the non-classical monocytes, such a correlation was not observed within any of the individual groups indicating that these markers might not be robustly associated with CSVD and VCI during HIV infection.

There are some limitations to our study. While PLWH were well matched to HIV uninfected participants for age, sex and Reynold’s cardiovascular risk score (70), a larger sample size and a more equal distribution of sex-at-birth would have been preferable. Unfortunately, the HIV population in the recruitment geographic area is heavily skewed towards males. Every effort was made to enroll female study participants which led to at least 25% female study participants in each cohort. Of notice, the cardiovascular risk score was comparable between the HIV+ and HIV- cohorts and also between HIV-CSVD+ group and HIV+CSVD+ group indicating that these groups share similar burden for development of cardiovascular disease in subsequent years. Although no specific treatment for CSVD is available, many of the traditional vascular risk factors such as diabetes, hypertension and hyperlipidemia are treatable. Therefore, stronger surveillance and prompt treatment should be in place for PLWH to prevent CSVD.

In conclusion, elevated levels of non-classical monocytes in PLWH may represent a common pathway to neuroinflammation and CSVD thus cognitive impairment. However, in depth mechanistic studies that incorporate inhibitors of platelet-monocyte interaction, for example inclacumab, anti-p-selectin antibody (71), or anti-platelet drugs such as P_2_Y_12_ inhibitors (72–74) in animal models of HIV infection are needed as further validation. In addition to its potential contribution to disease, level of non-classical monocytes may also be a useful biomarker to assess response to intended CSVD treatments or to monitor progression of CSVD. In this regard, evaluation of results from longitudinal follow up visits, which are currently ongoing, might provide valuable insights.

## MATERIALS AND METHODS

### Study design

Inclusion and exclusion criteria and all the study procedures are described in detail in our previous report (29). A total of 110 PLWH and equal number of age, sex-matched HIV uninfected individuals were enrolled. Demographic characteristics are described in **Table 1**. While the study participants are followed longitudinally for two more visits (at 18 and 36 months after enrollment), the data presented in this report is cross-sectional and only pertains to the baseline study visit. The study is still conducting longitudinal investigations.

**Table 1:** Demographic characteristics of the study participants. For age, Reynold’s risk score (RRS), CD4 counts, viral load and white matter hyperintensities (WMH) volumes, values are presented as mean (standard deviation). INSTI: integrase strand transfer inhibitor, NNRTI: non- nucleoside reverse transcriptase inhibitor, PI: protease inhibitor, DWMH: deep white mater hyperintensities; PVWMH: periventricular white matter hyperintensities, HAND: HIV associated neurological disorder, ANI: asymptomatic neurocognitive impairment, MND: mild neurocognitive impairment, HAD: HIV-associated dementia.

| Category | Variable | HIV-CSVD- | HIV-CSVD+ | HIV+ CSVD- | HIV+ CSVD+ |
| --- | --- | --- | --- | --- | --- |
| N |  | 40 | 68 | 30 | 81 |
| Age |  | 44.08 (15.31) | 54.75 (16.17) | 48.13 (13.09) | 54.62 (10.05) |
| Sex | Female | 11 | 20 | 8 | 20 |
|  | Male | 28 | 48 | 22 | 68 |
|  | Other | 1 | 0 | 0 | 0 |
| Race/<br>Ethnicity | Hispanic | 2 | 5 | 7 | 5 |
|  | Not Hispanic | 37 | 63 | 23 | 75 |
|  | White | 33 | 61 | 15 | 51 |
|  | Black | 3 | 7 | 12 | 26 |
|  | Other | 4 | 0 | 0 | 4 |
|  | Not Reported | 1 | 0 | 0 | 1 |
| Clinical | Diabetes Type II (Yes: No) | 0:40 | 0:68 | 1:29 | 10:71 |
|  | Hepatitis C | 0:40 | 1:67 | 2:28 | 7:74 |
|  | Hypertension | 10:30 | 18:50 | 8:22 | 29:52 |
|  | RRS | 4.025 (4.44) | 7.831 (7.173) | 4.233 (3.997) | 7.654 (6.751) |
| HIV<br>related | CD4 Count (Cells/mm<sup>3</sup>) | - | - | 692 (264.6) | 650 (325.2) |
|  | Viral Load (Copies/mL) | - | - | 4.7 (8.8) | 9.95 (19.4) |
|  | cART (INSTI) | - | - | 18 | 67 |
|  | cART (NNRTI) | - | - | 8 | 22 |
|  | cART (PI) | - | - | 4 | 15 |
| CSVD | DWMH | - | - | 65:03 | 79:02 |
|  | PVWMH | - | - | 23:45 | 42:39 |
|  | WMH Volume (cm<sup>2</sup>) | - | - | 0.088 (2.64) | 0.512 (0.808) |
| HAND<br>category | ANI | - | - | 11 | 44 |
|  | MND | - | - | 5 | 9 |
|  | HAD | - | - | - | 1 |

**Table 2:**
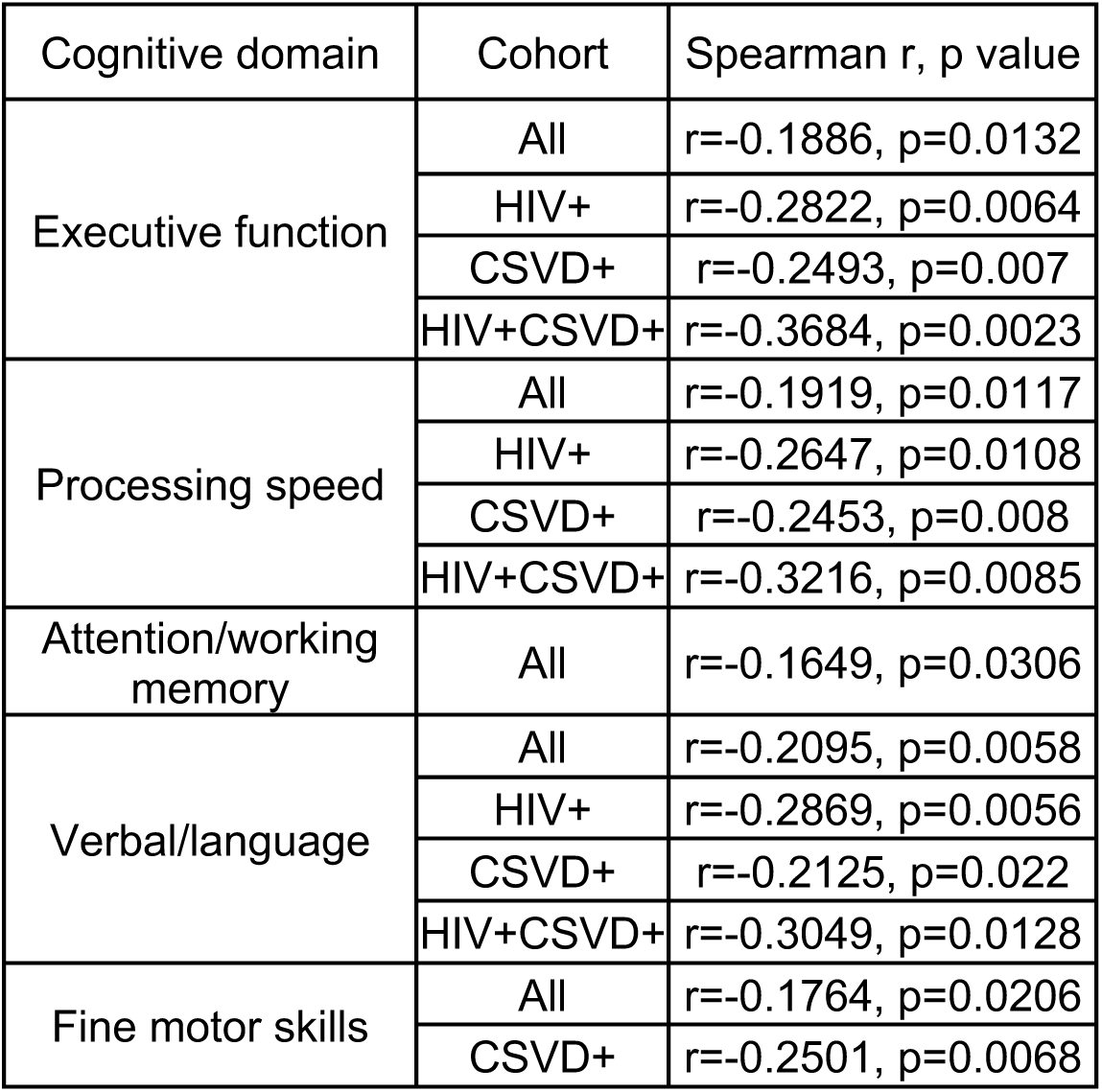
Correlation analysis between percentages of non-classical monocytes and individual cognitive domain scores. Only statistically significant association are shown.

| Cognitive domain | Cohort | Spearman r, p value |
| --- | --- | --- |
| Executive function | All | r=-0.1886, p=0.0132 |
|  | HIV+ | r=-0.2822, p=0.0064 |
|  | CSVD+ | r=-0.2493, p=0.007 |
|  | HIV+CSVD+ | r=-0.3684, p=0.0023 |
| Processing speed | All | r=-0.1919, p=0.0117 |
|  | HIV+ | r=-0.2647, p=0.0108 |
|  | CSVD+ | r=-0.2453, p=0.008 |
|  | HIV+CSVD+ | r=-0.3216, p=0.0085 |
| Attention/working memory | All | r=-0.1649, p=0.0306 |
| Verbal/language | All | r=-0.2095, p=0.0058 |
|  | HIV+ | r=-0.2869, p=0.0056 |
|  | CSVD+ | r=-0.2125, p=0.022 |
|  | HIV+CSVD+ | r=-0.3049, p=0.0128 |
| Fine motor skills | All | r=-0.1764, p=0.0206 |
|  | CSVD+ | r=-0.2501, p=0.0068 |

### Neuropsychological Battery

All cognitive testing was performed by staff trained and supervised by a clinical neuropsychologist. We administered a comprehensive neuropsychological battery that included tests of Attention and Working Memory (CalCAP), Processing Speed (Symbol Digit Modalities Test and Stroop Color Naming), Executive Function (Trail making Test Parts B, Stroop Interference task), Fine Motor skills (Grooved Pegboard, left and right hand); Verbal and visual learning (Rey Auditory Verbal Learning Test AVLT (Trials 1-5), Rey Complex Figure Test Immediate Recall), verbal and visual memory (Rey Auditory Verbal Learning Test RAVLT Delayed Recall, Rey Complex Figure Test Delayed Recall) and language (verbal fluency tests) at each visit. Premorbid intellectual functioning ability was estimated via WRAT-4 Reading at the baseline visit only.

Raw scores from each test were converted to z-scores using test manual norms. We created cognitive domain scores by averaging the z-scores of each test within each domain. We created a total cognitive score was by summing the six cognitive domain z-scores (Attention and Working Memory, Processing Speed, Executive Function, Fine Motor Skill, Verbal and Visual Learning, Verbal and Visual Memory and Language). We used Frascati criteria (75) to determine HAND diagnoses for each participant.

### MRI acquisition, analysis and CSVD diagnosis

Imaging was conducted on a Siemens 3T MAGNETOM PrismaFit whole body MRI scanner (Erlangen, Germany; software version VE11c) equipped with 64-channel phased array head coil. Detailed study protocols have already been reported by our group (29). Briefly, the following imaging was acquired: high-resolution 3D T1-weighted (T1w) anatomical images using a magnetization prepared rapid gradient echo (3D MPRAGE) sequence (inversion time -TI=926 ms; repetition time/echo time -TR/TE=1840/2.45 ms; image resolution = 1 × 1 × 1 mm^3^); 2D T2-weighted (T2w) images using a turbo spin echo sequence (TR/TE = 6000/100 ms; image resolution = 0.5 × 0.5 × 5 mm^3^); 3D fluid attenuated inversion recovery (FLAIR) image (TI = 1800 ms; TR/TE = 5000/100 ms; resolution = 1 × 1 × 1 mm^3^); and 3D T2*-weighted images using a multi-echo gradient echo (MEGE) sequence with bi-polar readout (TR/TE 1st echo = 84/5.43 ms; number of echoes = 8; image resolution = 0.94 × 0.94 × 2.0 mm^3^). The T1w, T2w, T2*w and FLAIR images were used to evaluate CSVD burden by an experienced neuroradiologist using the Fazekas score (76). Finally, VolBrain (77) was used to quantitatively measure the total WMH burden using the T1w and FLAIR images.

### Whole blood collection and processing

Approximately, 40 ml of whole blood was collected in Acid Citrate Dextrose (ACD) vacutainers, incubated at room temperature with slow shaking and processed within 2 hours of collection. Plasma was isolated by centrifugation at 1000 X g for 10 minutes at room temperature. Plasma was aliquoted and cryopreserved at -80 °C and used to ELISA analyses mentioned below. 1ml of whole blood was processed by flow cytometry to measure the levels of activated platelets (i.e., platelets expressing CD62p), circulating platelet-monocyte complexes (PMCs) and expression of various monocyte activation markers such as c-c chemokine receptor 2 (CCR2), CD40, p-selectin glycoprotein ligand (PSGL-1), tumor necrosis factor receptor 2 (TNFR2) and tissue factor (TF).

### Flow cytometric analysis of whole blood

Flow cytometric analysis of platelet-monocyte complexes and platelet activation was performed as described previously (22, 29, 41). Briefly, within two hours of draw, whole blood was fixed with paraformaldehyde (PFA) followed by RBC lysis using ACK lysis buffer. The cells were then washed and stained with titrated amounts of antibodies against anti-CD14 PE (BD Biosciences # 555398; 10 ul), anti-CD16 PE Cy7 (BD Biosciences 557744; 3 ul), anti-CD61AF647 (Biolegend # 336408; 3 ul), anti-PSGL-1 FITC (R&D Systems # FAB9961G; 1.5 ul), anti-CD40 FITC BD Biosciences. # 555598; 10 ul), anti-CCR2 FITC (R&D Systems # FAB151G; 1.5 ul), anti-CD62P FITC (BD Biosciences # 555523; 5 ul) and anti-TNFR2 FITC (Miltenyi Biotech #130-107-743; 1 ul) for 30 minutes at room temperature in dark. The cells were washed and acquired using Accuri C6 flow cytometer. 75,000 gated leukocytes were acquired based on forward and side scatter per tube. Data was analyzed using Flow Jo software version 10.4.2. Unstained cells, cell stained with anti-CD14 and anti-CD16 were used to gate on three subsets of monocytes, classical monocytes (CD14+ CD16-), intermediate monocytes (CD14+CD16+) and non-classical monocytes (CD14- CD16+). Among these cells, those that expressed CD61, a platelet marker, were categorized as PMCs. Expression of CCR2, CD40, CD62p, TNFR2 and TF was measured on all monocytes and also on PMCs. Further, 10000 platelet events were acquired based on size beads (0.9 to 3 um) and expression of CD61. Platelet activation was measured by expression of CD62p.

### ELISAs

Plasma samples were used to conduct ELISA assays for ICAM (# DY720-05, R and D systems, MN, USA), VCAM (# DY809, R and D systems, MN, USA), sCD14 (# DY383, R and D systems, MN, USA), CD163 (# DY1607, R and D systems, MN, USA), Osteoprotegerin (# DY805, R and D systems, MN, USA) and lipoprotein associated phospholipase A2 (LpPLA2) (# ab235643, Abcam, Cambridge, UK) as per manufacturer’s instructions.

### Statistical analysis

Descriptive characteristics of study participants are as shown in Table 1. Percentages and frequencies for discrete variables were reported as well as the means and standard deviations for continuous characteristics. D’Agostino & Pearson test was performed to analyze the normality of the data. If the normality assumption was rejected, an appropriate nonparametric test was performed; otherwise, a parametric test was used. Mann-Whitney U test (nonparametric) or two sample Welch t-test (parametric) was used to perform two group comparisons (HIV+ and HIV- groups). Four group comparisons (HIV-CSVD-, HIV-CSVD+, HIV+CSVD-, HIV+CSVD+ groups) were done by Kruskal-Wallis test followed by Dunn’s multiple comparisons test (nonparametric) or by one-way ANOVA F-test followed by Tukey’s multiple comparisons test (parametric). Two group comparisons between PMCs and non-PMCs were performed by the Wilcoxon signed-rank test (nonparametric) or paired t-test (parametric). All correlation analyses were done using Spearman’s rank correlation test. A p value of less than 0.05 was considered significant. Error bars represent mean ± standard deviation. All statistical analyses were performed with GraphPad Prism 9.4.1 (GraphPad Software, Inc., San Diego, CA). **Study approval**

The study was approved by University of Rochester Research Subjects Review Board. Written informed consents were obtained from all study participants before enrollment into the study as per the Declaration of Helsinki.

## Data Availability

All data produced in the present study are available upon reasonable request to the authors

## ACKNOWLEDGEMENTS

The authors would like to acknowledge funding sources NIH R01s AG054328 (to GS and SBM), MH118020 (to GS). We would like to thank Ann Miller and the whole team at Clinical Research Center, URMC for help with the blood draws. We would like to acknowledge the resources and colleagues at the Flow cytometry core at URMC. Our sincere thanks to Jill Guary, Teresa Oh and Allison Preteroti for help with study recruitment, and Kyle Murray, Yuchuan Zhuang, Arun Venkataraman, Nhat Hoang, Alan Finkelstein and Abrar Faiyaz for support with imaging processing.

## AUTHOR CONTRIBUTIONS

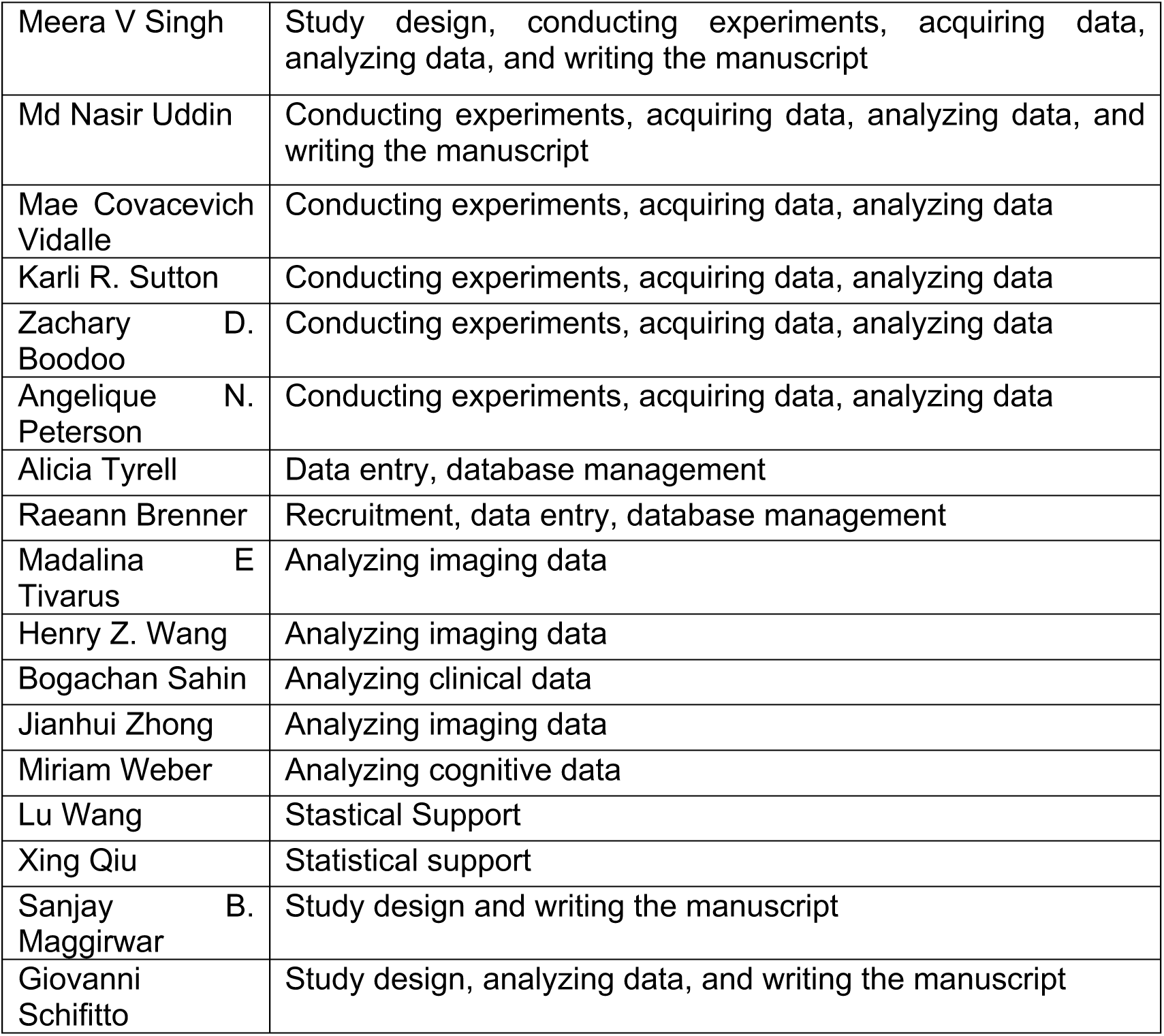

