## Supplementary figures 1 to 3 for "Role of non-classical monocytes in HIV-associated vascular cognitive impairment"

**Supplementary figure 1:** **Flow cytometric analysis of interaction between classical or intermediate monocytes and platelets.** Percentages of platelet-monocyte complexes (PMCs) among classical monocytes **A.** based on HIV status (HIV- n=81, HIV+ n=94), **B.** HIV and CSVD status (HIV-CSVD- n=31, HIV-CSVD+ n=50, HIV+CSVD- n=26, HIV+CSVD+ n=68); among intermediate monocytes **C.** by HIV status, (HIV- n=81, HIV+ n=94), **D.** HIV and CSVD status, (HIV-CSVD- n=31, HIV-CSVD+ n=50, HIV+CSVD- n=26, HIV+CSVD+ n=68). Light blue data points represent HIV-CSVD- group, dark blue represent HIV-CSVD+ group, orange represent HIV+CSVD- group and red represent HIV+CSVD+ group. Two group comparisons by Mann-Whitney U test, four group comparisons by Kruskal-Wallis test followed by Dunn’s multiple comparisons test. * p<0.05, ** p<0.01, *** p<0.001.

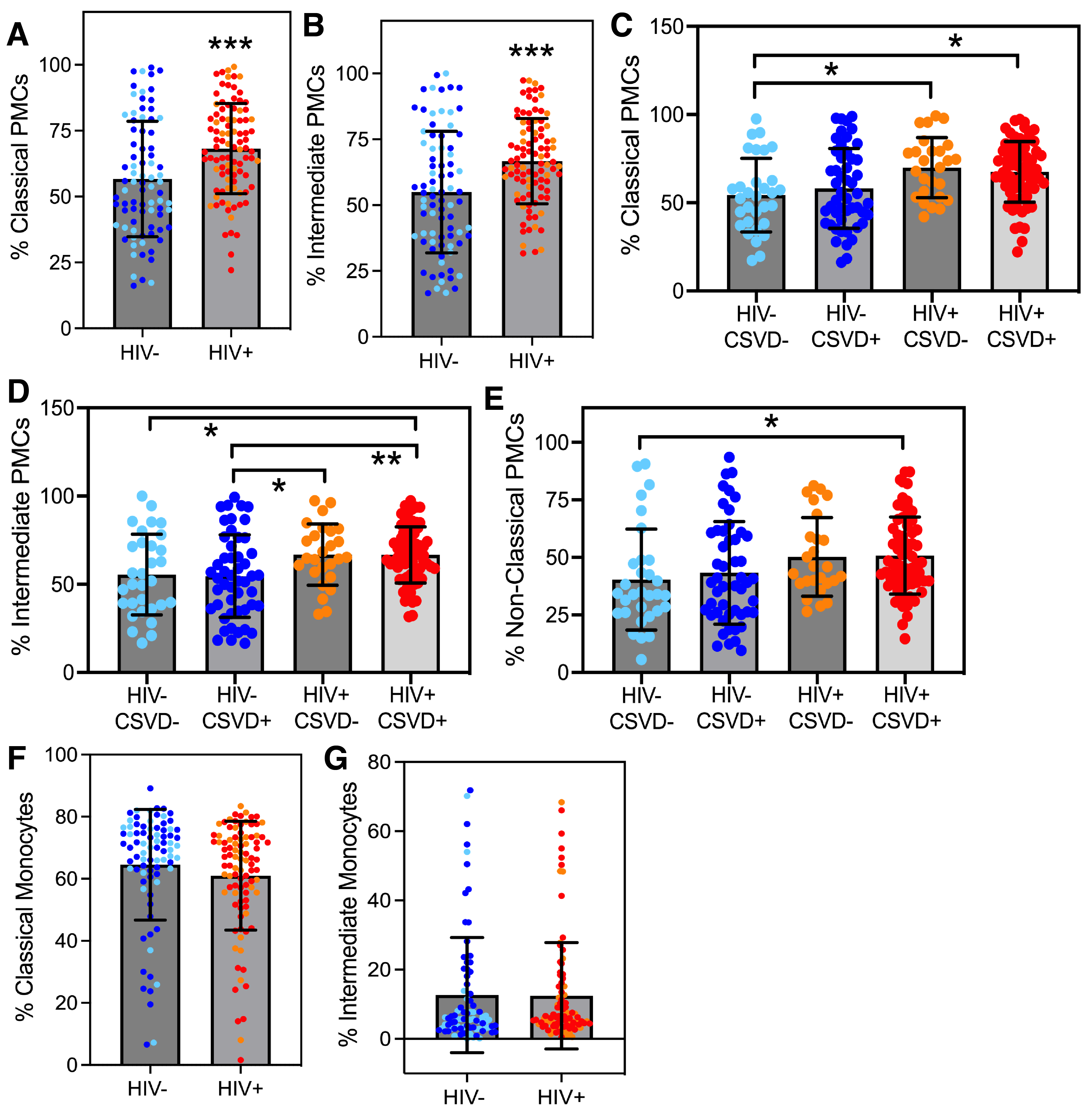

**Supplementary figure 2: Levels of monocyte activation markers among classical and intermediate monocyte subsets.** Levels of **A.** classical (HIV- n=80, HIV+ n=93) and **B.** intermediate monocytes expressing tissue factor (HIV- n=80, HIV+ n=93), **C.** classical monocytes expressing CD40 (HIV- n=81, HIV+ n=94), irrespective of whether the cells are complexed to platelets or not. Two group comparisons by Mann-Whitney U test, * p<0.05, ** p<0.01. Comparison between percentages of platelet-complexed (PMCs) and non-complexed (non-PMCs) classical monocytes for expression levels of **D.** Tissue Factor (TF, n=173), **E.** CD40 (n=175), **F.** Tissue necrosis factor receptor 2 (TNFR2, n=175), **G.** Comparison between percentages of platelet-complexed (PMCs) and non-complexed (non-PMCs) intermediate monocytes for expression levels of TNFR2 (n=175). Light blue data points represent HIV-CSVD- group, dark blue represent HIV-CSVD+ group, orange represent HIV+CSVD- group and red represent HIV+CSVD+ group. Two group comparisons between PMCs and non-PMCs by Wilcoxon signed-rank test, * p<0.05, **** p<0.0001.

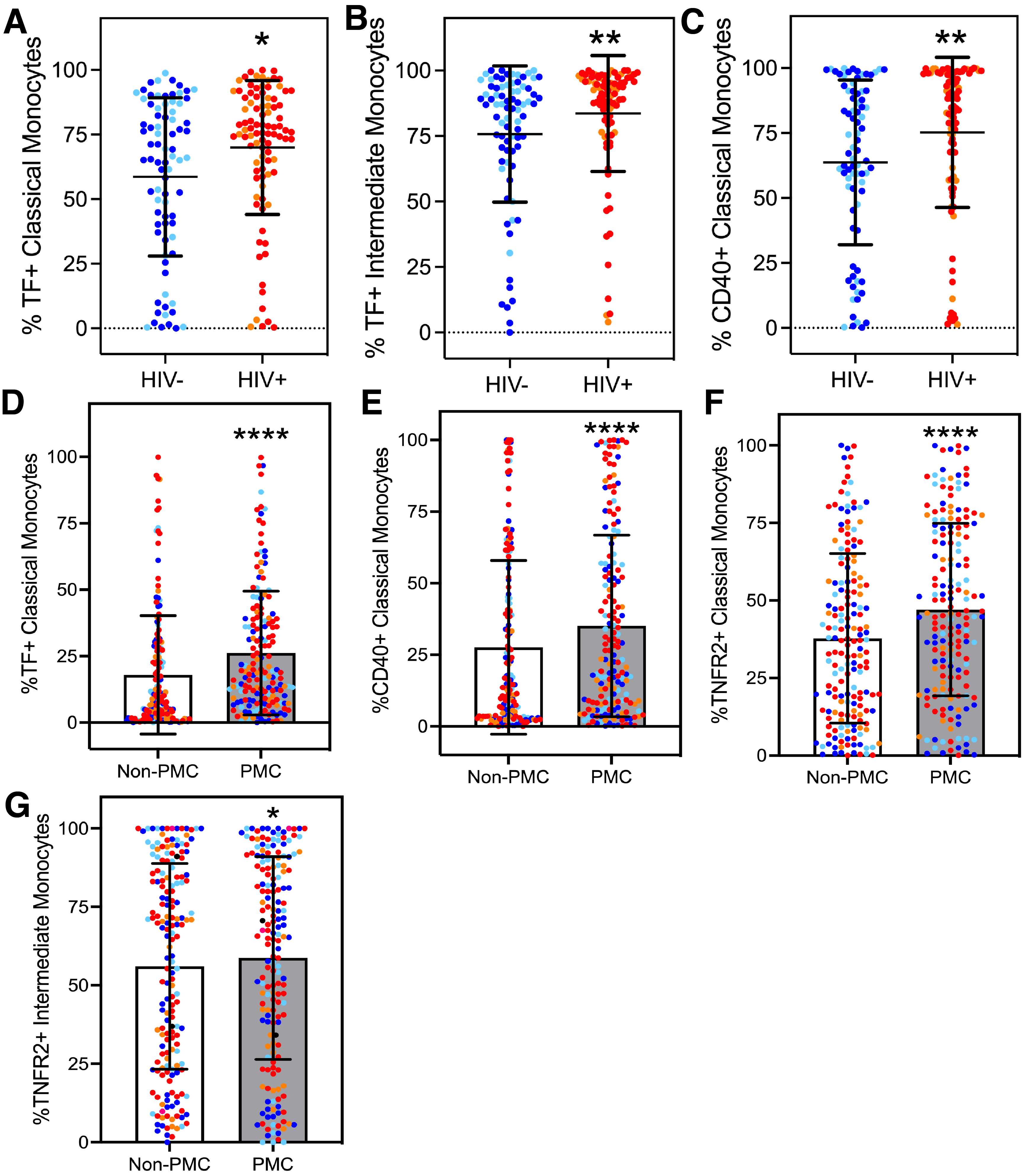

**Supplementary figure 3: Soluble markers of endothelial and monocyte activation and correlation with cognitive performance.** Plasma levels of **A.** vascular adhesion molecule (VCAM, HIV-CSVD- n=37, HIV-CSVD+ n=65, HIV+CSVD- n=26, HIV+CSVD+ n=78), **B.** LpPLA2 (HIV-CSVD- n=39, HIV-CSVD+ n=65, HIV+CSVD- n=28, HIV+CSVD+ n=79), **C.** Osteoprotegerin (HIV-CSVD- n=37, HIV-CSVD+ n=65, HIV+CSVD- n=27, HIV+CSVD+ n=77) based on HIV and CSVD status, CD163 based on **D.** HIV status (HIV- n=103, HIV+ n=105), E. HIV and CSVD status (HIV-CSVD- n=40, HIV-CSVD+ n=63, HIV+CSVD- n=27, HIV+CSVD+ n=78). Correlation analysis between percentages of non-classical monocytes (irrespective of whether the cells are complexed to platelets or not) and total cognitive scores within **F.** HIV- study group (n=80, r=0.0307) and **G.** HIV+CSVD- group (n=26, r=-0.0906). Light blue data points represent HIV-CSVD- group, dark blue represent HIV-CSVD+ group, orange represent HIV+CSVD- group and red represent HIV+CSVD+ group. Four group comparisons by Kruskal-Wallis test followed by Dunn’s multiple comparisons test. Correlation analysis by Spearman’s rank correlation test. * p<0.05.

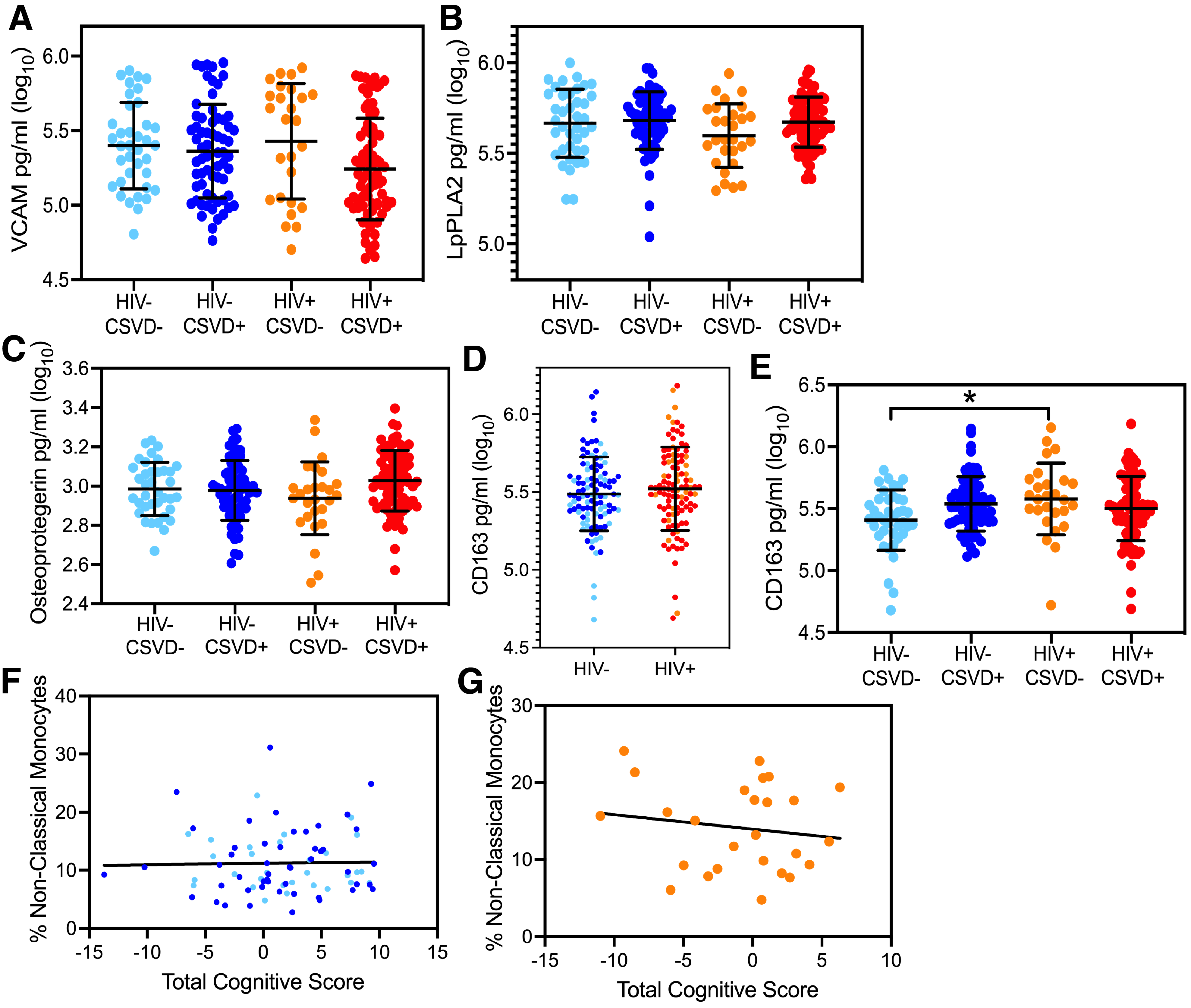
